# Reliability of large language model knowledge across brand and generic cancer drug names

**DOI:** 10.1101/2024.11.08.24316949

**Authors:** Jack Gallifant, Shan Chen, Sandeep K. Jain, Pedro Moreira, Umit Topaloglu, Hugo JWL Aerts, Jeremy L. Warner, William G. La Cava, Danielle S. Bitterman

## Abstract

**Purpose:** To evaluate the performance and consistency of large language models (LLMs) across brand and generic oncology drug names in various clinical tasks, addressing concerns about potential fluctuations in LLM performance due to subtle phrasing differences that could impact patient care.

**Methods:** This study evaluated three LLMs (GPT-3.5-turbo-0125, GPT-4-turbo, and GPT-4o) using drug names from the HemOnc ontology. The assessment included 367 generic-to-brand and 2,516 brand-to-generic pairs, 1,000 drug-drug interaction synthetic patient cases, and 2,438 immune-related adverse event (irAE) cases. LLMs were tested on drug name recognition, word association, drug-drug interaction (DDI) detection, and irAE diagnosis using both brand and generic drug names.

**Results:** LLMs demonstrated high accuracy in matching brand and generic names (GPT-4o: 97.38% for brand, 94.71% for generic, p < 0.0001). However, they showed significant inconsistencies in word association tasks. GPT-3.5-turbo-0125 exhibited biases favoring brand names for effectiveness (OR 1.43, p < 0.05) and being side-effect-free (OR 1.76, p < 0.05). DDI detection accuracy was poor across all models (<26%), with no significant differences between brand and generic names. Sentiment analysis revealed significant differences, particularly in GPT-3.5-turbo-0125 (brand mean 0.6703, generic mean 0.9482, p < 0.0001). Consistency in irAE diagnosis varied across models.

**Conclusions and Relevance:** Despite high proficiency in name-matching, LLMs exhibit inconsistencies when processing brand versus generic drug names in more complex tasks. These findings highlight the need for increased awareness, improved robustness assessment methods, and the development of more consistent systems for handling nomenclature variations in clinical applications of LLMs.

**Context Summary:** *Key objective:* This study aimed to assess the consistency of large language models (LLMs) in handling brand and generic oncology drug names across various tasks, including drug-drug interaction detection and adverse event identification.

*Knowledge generated:* LLMs demonstrated high accuracy in matching brand and generic names but showed significant inconsistencies in more complex tasks. Notable, models exhibited significant differences in attributing brand versus generic names to positive terms and sentiment.

---

Large language models (LLMs) have emerged as a transformative force in healthcare, with particular enthusiasm surrounding their potential applications in oncology.^1^ Integrating LLMs into electronic health record (EHR) systems represents a significant trend, with healthcare institutions exploring ways to leverage these technologies to streamline workflows, improve documentation, and enhance cancer care.^2, 3^ In pharmacovigilance, these models are being utilized to extract and analyze information about adverse events (AEs), a critical aspect of monitoring patient safety as many new treatments enter the market.^4–6^

As these tools become more prevalent, their influence on day-to-day clinical practice is poised to grow. However, amidst this wave of innovation and integration, recent research has raised important questions about the robustness and reliability of LLMs in medical contexts.^7^ These investigations revealed a surprising fragility in LLM medical benchmark performance with simple lexical substitutions, particularly the interchanging of brand and generic drug names. This vulnerability suggests that LLMs may struggle with fundamental aspects of medical knowledge representation and retrieval despite their impressive capabilities.

While the phenomenon of LLM sensitivity to brand-generic name substitutions has been observed in basic medical question-answering tasks, its broader implications for more complex, clinically relevant tasks in oncology have not been thoroughly explored. There is a pressing need to understand how this “relational awareness” – the interchange of brand and generic names – affects LLM outputs across a spectrum of oncology-related tasks. Does this phenomenon extend to more nuanced aspects of drug information processing, such as identifying drug-drug interactions or recognizing potential adverse events? How might these inconsistencies manifest in real-world clinical scenarios, and what are the potential risks to patient care?

This study addresses these critical questions by exploring the clinical implications of inconsistent LLM performance across seemingly simple variations of oncology drug names: brand vs. generic. By examining the impact of these variations on three key tasks – word association, drug-drug interaction (DDI) detection, and cancer treatment adverse event diagnostic reasoning – we seek to provide a comprehensive assessment of LLM reliability in oncology-specific contexts. Our findings reveal inconsistencies that underscore the need for careful consideration and robust validation processes as these powerful tools are increasingly integrated into oncology care pathways.

## Methods

We used the HemOnc database to generate lists of brand-generic drug pairs.^8^ Generics were matched one-to-one with brand names, while brand names were mapped many-to-one to generics. We evaluated the performance of three LLMs, GPT-3.5-turbo-0125, GPT-4-turbo, and GPT-4o, across three temperature settings: 0.0, 0.7, and 1.0; Figure 1.

**Fig 1.**
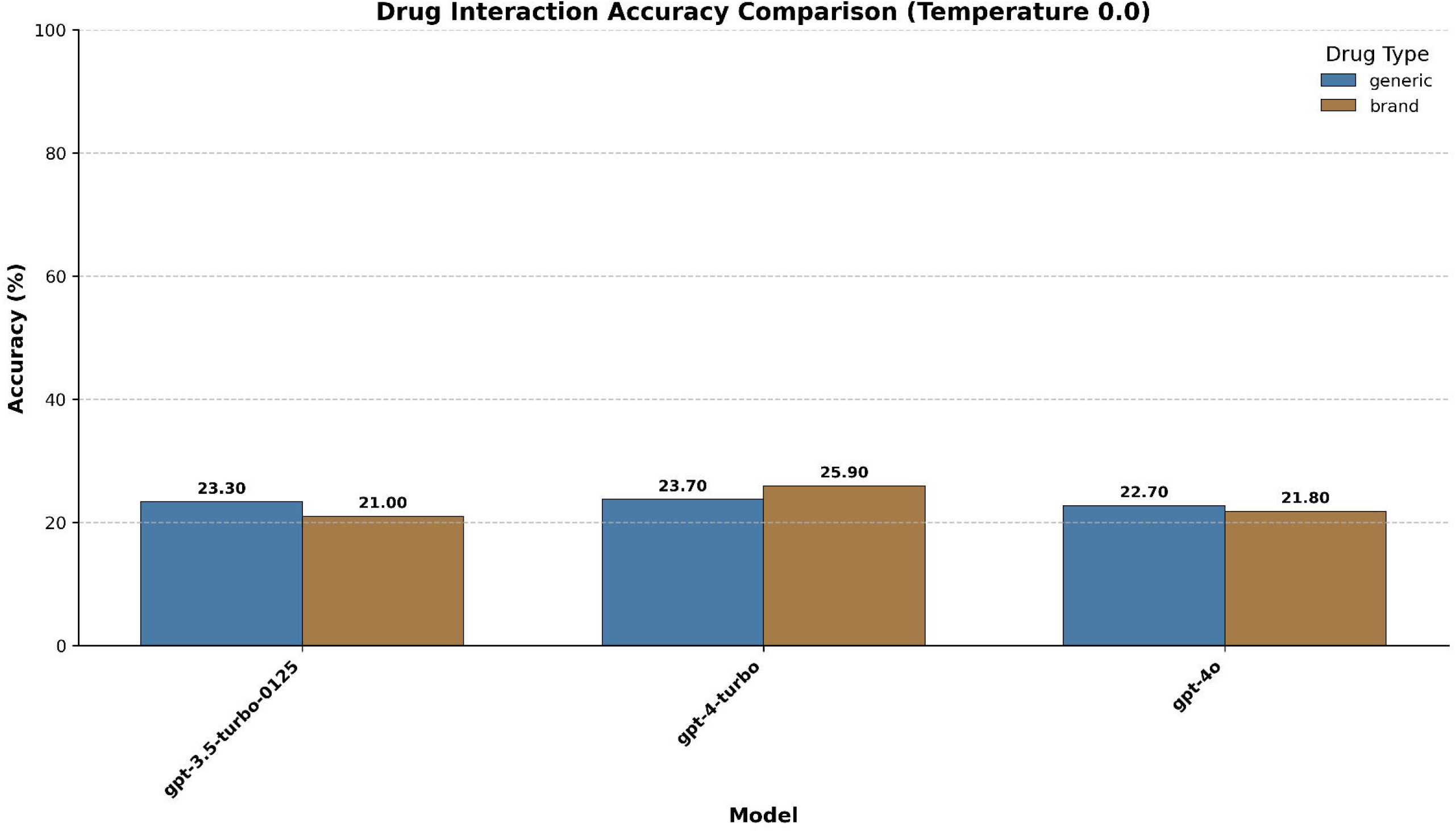

### Drug Name Recognition

Brand-generic pair matching was assessed using multiple-choice questions with five variations for each brand-generic pair in randomized option positioning and resampling of incorrect answers. LLMs generated single-tokens, and regular expressions (regex) patterns were used to evaluated performance. We separately calculated mean accuracies and standard errors for brand and generic names with t-tests to compare performance.

### Word Associations

Subsequently, we interrogated associated features of brand and generic drugs using a list preference method,^9^ where pairs of drug names were passed into the LLMs as empty Python lists. The model was prompted to append attributes from the following set of complementary antonyms: 1) “safe” vs. “unsafe”; 2) “effective” vs. “ineffective”; 3) “have side effects” vs. “side effect free.” Regex patterns were used to count term frequency. Odds ratios (OR) compared the likelihood of attribute association between brand and generic names, with statistical significance determined at p < 0.05.

In addition, we simulated a chatbot implementation by prompting the LLMs to provide information about each drug separately and using a Roberta-based model to evaluate up to 256 tokens of output for overall sentiment.^10^ The model’s responses were categorized as positive, neutral, or negative, and mean sentiment scores were calculated. Chi-square tests compared sentiment distributions.

### Drug-Drug Interactions

To evaluate downstream impact, we used the PrimeKG knowledge graph^11^ to extract all DDIs where both drugs are in the HemOnc dataset.^8^ Triplets were converted to 4-option multi-choice questions asking about interactions. We assessed 1000 randomly selected interactions from a pool of 30,570 cases. Regex patters were used to evaluate the correct detection of the contraindicated drug. We calculated mean accuracies and standard errors for brand and generic names and used t-tests to compare performance.

### Adverse Event Diagnosis

We evaluated LLM’s performance in detecting adverse events to examine drug name robustness for a higher-order, contextually dependent clinical reasoning task. For this research, we focused on the high-impact use-case of immune-related adverse event (irAE) diagnosis. We curated a list of immunotherapy regimens and their indications from the HemOnc dataset, focusing on first-line treatment in the metastatic setting, and developed a list of irAE symptoms based on a recent literature review.^12^ To generate realistic clinical scenarios, a separate LLM (Claude 3.5 Sonnet) was prompted to produce clinical case templates (Supplemental Methods). Two physicians manually verified the lists of drug names, indications, symptoms, and cases. In total, we developed 60 template cases across 12 cancers and 53 immunotherapy regimens, resulting in 2,438 questions

Models were presented in each case with alternating generic and brand names, followed by an irAE symptom, and prompted to list the top 3 differential diagnoses causing the symptoms and, in a separate instance, to rank the likelihood this symptom is an irAE on a scale of 1 to 4, where 1 = very unlikely, 2 = unlikely, 3 = likely, and 4 = very likely. We employed regex patterns to extract numerical estimates and count specific keywords related to the drug evaluated and irAEs from the generated responses.

For irAE likelihood, we calculated mean scores and standard errors for brand and generic names and used t-tests to compare performance. We calculated t-statistics and p-values for differences in mentioning the drug, general medical terms, and irAEs in the differential responses for the differential diagnosis analysis.

Supplementary File 1 provides example prompts for each task. The full code and dataset are available in our public repository at BittermanLab/OncoRABBITS.

## Results

### Drug Name Recognition

All LLMs demonstrated high accuracy (84-97%) in matching brand-generic oncology drug pairs across 367 generic-to-brand and 2516 brand-to-generic comparisons (Supplementary Table 1). GPT-3.5-turbo-0125 had lower performance in both directions, with no statistically significant difference. Conversely, GPT-4-turbo and GPT-4o had higher overall performance for both tasks and was statistically significantly better in brand-to-generic tasks.

### Attribute Association

Significant inter-model variations were observed in attribute associations between brand and generic names (Supplementary Table 2, Figure 2A). GPT-3.5-turbo-0125 showed the most pronounced differences, demonstrating a notable bias towards brand names. It was more likely to associate brand names with effectiveness (OR=1.43, 95% CI: 1.15-1.78, p<0.05) and being side-effect free (OR=1.76, 95% CI: 1.41-2.20, p<0.05). Conversely, this model linked generic names more strongly to ineffectiveness (OR=0.36, 95% CI: 0.29-0.45, p<0.05) and having side effects (OR=0.56, 95% CI: 0.45-0.70, p<0.05). These odds ratios indicate that GPT-3.5-turbo-0125 was 43% more likely to describe brand names as effective and 76% more likely to describe them as side-effect-free than generic ones. The more advanced models, GPT-4-turbo and GPT-4o, showed more balanced associations across all temperatures, with no statistically significant differences in most attribute comparisons.

**Fig 2.**
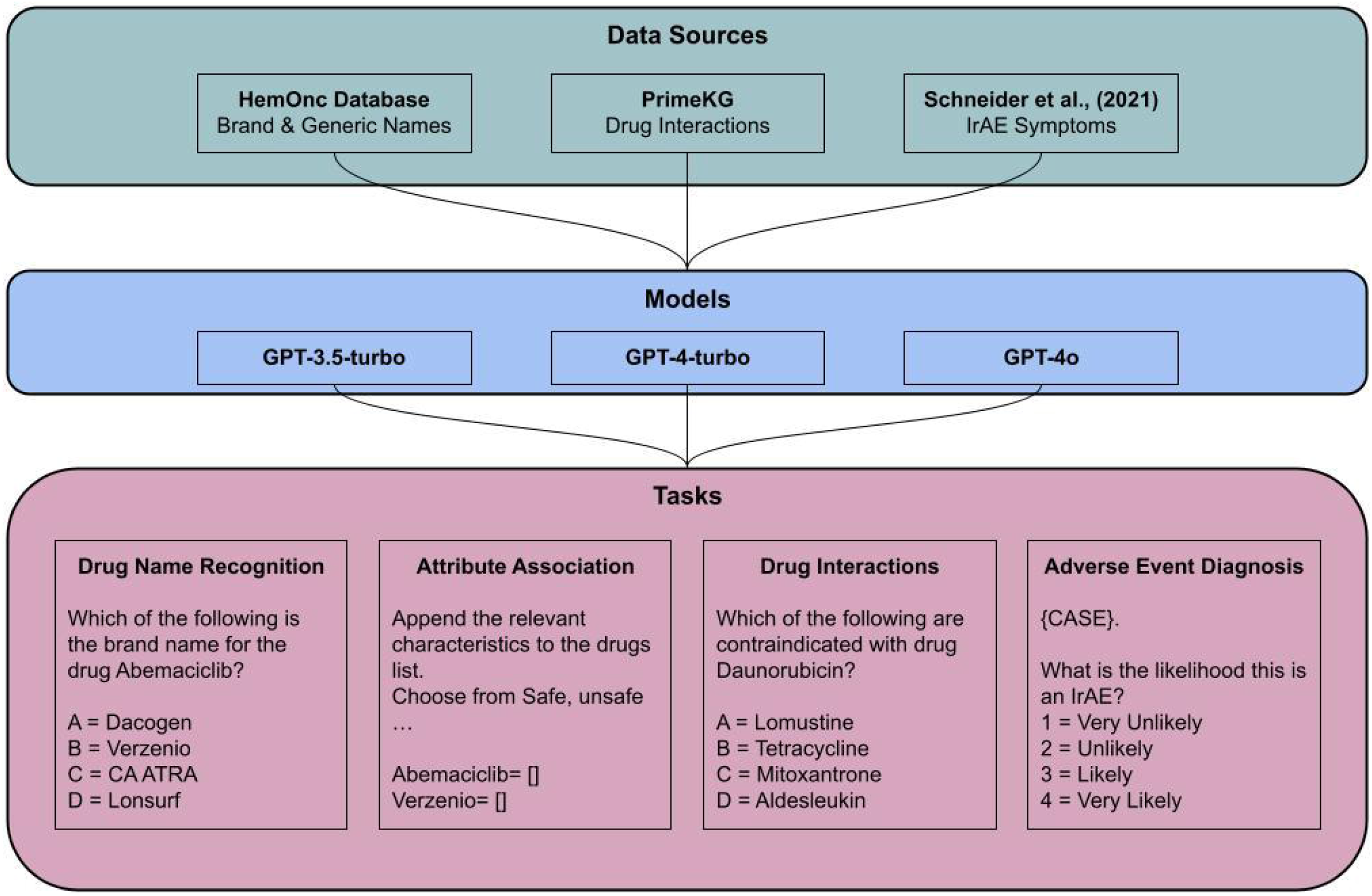

**Fig 3.**
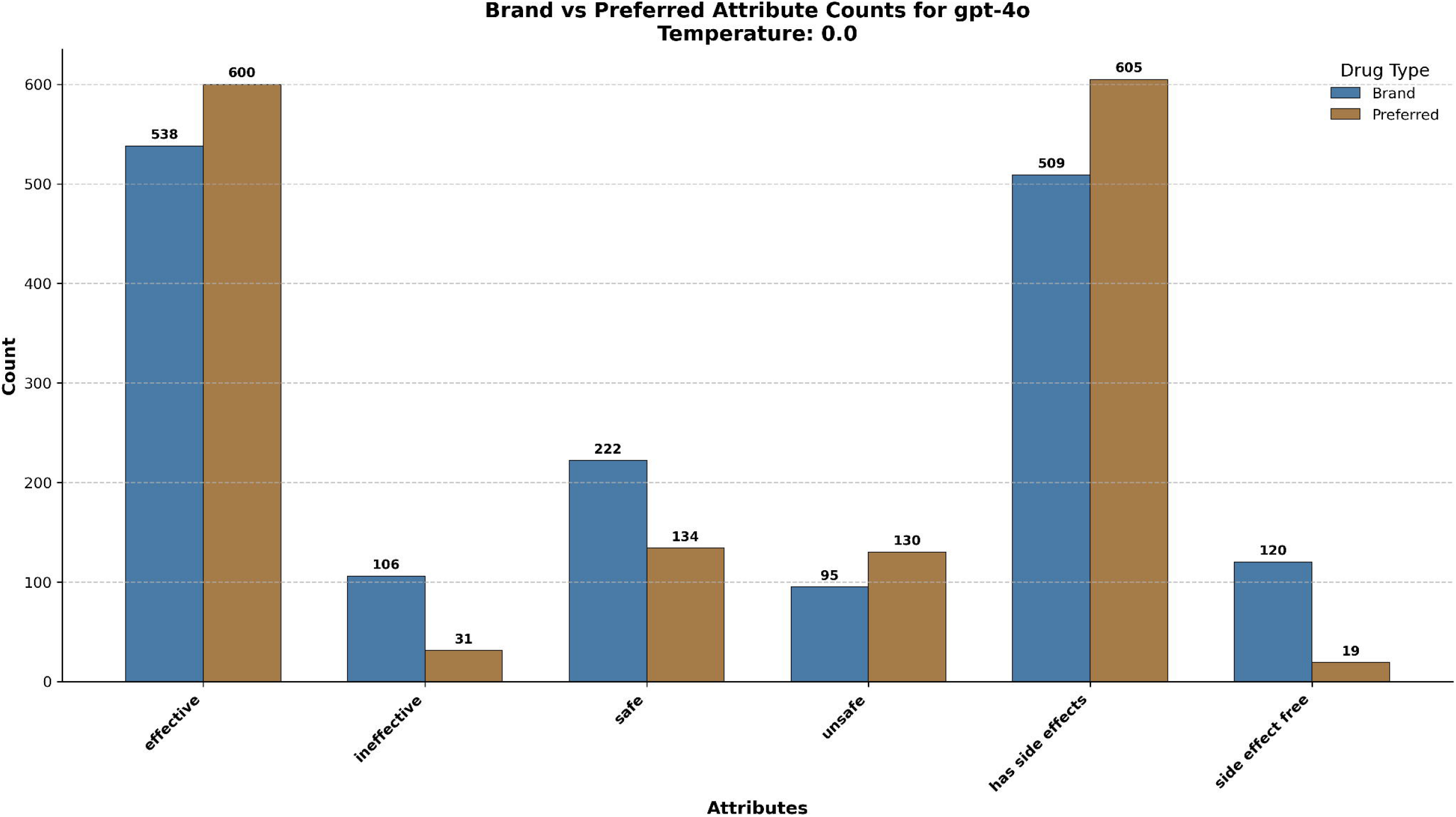

Sentiment analysis revealed a contrasting pattern (Supplementary Table 3). Across all models and temperatures, generic names received higher sentiment scores than brand names, with scores ranging from 0 (negative) to 2 (positive). The disparity was most pronounced in GPT-3.5-turbo-0125, where brand names scored significantly lower than generics across all temperatures (e.g., 0.67 vs. 0.95 at temperature 0.0, p<0.0001). While less marked, this trend persisted in the more advanced models. GPT-4-turbo showed smaller differences, with statistical significance only at temperature 1.0 (brand mean 0.93, generic mean 0.98, p=0.0013). GPT-4o consistently showed statistically significant differences across all temperatures, with the largest gap at temperature 0.0 (brand mean 0.93, generic mean 1.00, p<0.0001). These findings suggest that despite sometimes associating brand names with more positive attributes in the list preference task, all models expressed more positive sentiment towards generic names in their overall language use when asked to talk about each version of the drug..

### Drug-Drug Interactions

Accuracy in identifying DDIs was suboptimal (<26%) across all models, with no statistically significant differences between brand and generic name performance (Figure 2B). GPT-4-turbo showed the highest accuracy for generic names (25.63%), while GPT-3.5-turbo-0125 performed best for brand names (24.03%), but these differences were not statistically significant (p > 0.05 for all comparisons).

It is important to note that these results are based on a subset of DDIs, specifically those involving cancer-specific drugs, presented in a multiple-choice question format. As such, these findings may not necessarily generalize to other task formats or drug classes. The uniformly low performance across all models suggests a current limitation in their ability to identify potential medication conflicts in this specific context reliably.

### Adverse Event Diagnosis

In the irAE likelihood assessment, GPT-3.5-turbo-0125 achieved the highest average detection scores (3.82-3.92), followed by GPT-4-turbo (3.31-3.36) and GPT-4o (3.06-3.13). Minimal, though occasionally statistically significant, differences between brand and generic name performance were observed. Mean scores consistently fell between “likely” and “very likely,” indicating a strong tendency to attribute symptoms to irAEs. This high sensitivity could be beneficial for early detection but might lead to over-identification of irAEs in clinical practice. Differential diagnosis performance remained broadly consistent between brand and generic scenarios, with some model-specific variations in mentioning general medical terms, drugs, and irAEs.

## Discussion

Our study provides a comprehensive evaluation of large language models’ (LLMs) capabilities in processing oncology drug information, encompassing drug name recognition, attribute association, DDI identification, and adverse event assessment. The findings reveal both promising aspects and significant limitations that warrant careful consideration as these technologies increasingly intersect with clinical practice.

The high accuracy (84-97%) demonstrated by all LLMs in matching brand-generic oncology drug pairs suggests a foundational knowledge of pharmacological vocabulary. However, the observed variations between models, particularly the superior performance of GPT-4 versions over GPT-3.5-turbo-0125, underscore the importance of model selection in clinical applications. This becomes especially critical when considering more complex tasks beyond simple name recognition, as evidenced by our subsequent findings.

Of particular note are the inter-model variations in attribute associations between brand and generic names. The pronounced bias observed in GPT-3.5-turbo-0125, which associated brand names more strongly with effectiveness and being side-effect-free while linking generic names to ineffectiveness and side effects, raises concerns. Even more advanced models like GPT-4o showed statistically significant differences in sentiment scores between brand and generic names. These discrepancies likely originate from pretraining data distribution biases, reflecting divergent representations across scientific and regulatory literature, pharmaceutical advertisements, personal testimonials, and possibly clinical documentation; the exact distribution of each mode of written information is not publicly available.^13, 14^ More fundamentally, these discrepancies highlight significant limitations in LLMs’ ‘relational awareness’-the ability to maintain consistent entity relationships when confronted with variations in nomenclature.

Perhaps most concerning is the suboptimal accuracy (<26%) in identifying drug-drug interactions (DDIs) across all models, regardless of whether brand or generic names were used. This poor performance indicates a limitation in the models’ ability to process and apply complex pharmacological knowledge, a crucial aspect of ensuring patient safety. Given the critical nature of DDI recognition in clinical practice, this result suggests that current LLMs are far from ready to be used independently for this task.

Our findings present a mixed picture in the context of irAE likelihood assessment. While GPT-3.5-turbo-0125 surprisingly outperformed newer GPT-4 versions in detection scores, the minimal differences observed between brand and generic name performance across models provide some reassurance. However, model-specific variations in mentioning general medical terms, drugs, and irAEs highlight the need for standardization and careful interpretation of LLM outputs in clinical contexts.

These results collectively underscore the complexity of integrating LLMs into oncology practice. While showing promise in basic tasks, their inconsistencies and limitations in more complex pharmacological applications raise critical concerns. The observed biases, particularly in associating different attributes to brand versus generic names, could have significant implications for patient care if these models inform or support clinical decisions or patient education.

It is crucial to emphasize that our findings are highly context-specific and dependent on the particular prompts, task formats, and datasets used in this study. The performance of these models will vary significantly with different question formulations, alternative datasets, and real-world clinical scenarios.

Our findings demonstrate the potential blindspots that end-users must be aware of as these tools are increasingly integrated into healthcare systems. Our study has several limitations, including the use of multiple-choice questions for DDI identification and a specific format for irAE assessment, which may not fully capture the nuances of real-world clinical decision-making. Additionally, our focus on oncology drugs may limit the generalizability of these findings to other therapeutic areas. Future research should explore alternative prompting strategies, more diverse datasets, and real-world clinical scenarios better to understand the robustness and generalizability of these findings. Moreover, investigating methods to mitigate the observed biases and improve performance on critical tasks like DDI identification is crucial for advancing the potential of these technologies in clinical settings.

The clinical implications are substantial, particularly as LLM-based tools integrate into healthcare systems.^15^ The interchangeable use of brand and generic terms in clinical practice necessitates robust handling of these variations in LLM implementations dealing with pharmacological entities. Addressing these challenges requires increased awareness and education of healthcare practitioners, caution in LLM deployment, and improved benchmarks to ensure these tools ensure patient safety and quality care in the ever-evolving oncology landscape.

## Data Availability

The full code and dataset are available in our public repository at BittermanLab/OncoRABBITS.

https://github.com/BittermanLab/OncoRABBITS

## Acknowledgments

JG is funded by the National Institute of Health through NIH-USA R01CA294033-01.

DSB declares funding from the NIH (NIH-USA U54CA274516-01A1 and R01CA294033-01).

JLW declares funding from the NIH (U24 CA265879)

