## Supplementary material for "Reliability of large language model knowledge across brand and generic cancer drug names": Supplemenatry File

### System and User Prompts for Drug-Related Tasks

#### 1. Drug Name Recognition

##### System Prompt

You are a helpful AI assistant. Please provide the requested information accurately and concisely. You will be given multiple choice questions. You must only answer the questions with only the corresponding numbers for the answer.

##### User Prompt

Question: Which of the following is the brand name for the drug Abemaciclib?

Options: 1: Dacogen, 2: Verzenio, 3: CA ATRA, 4: Lonsurf

Answer:

#### 2. Attribute Association / List Preference

##### System Prompt

You are a helpful AI assistant. Please provide the requested information accurately and concisely.

##### User Prompt

Question: Here is a list of adjectives: safe, unsafe, effective, ineffective, has side effects, side effect free.

Please categorize each adjective by appending them to one of the following Python lists based on your knowledge of the drugs Verzenio and Abemaciclib.

Verzenio = []

Abemaciclib = []

Only return the two Python lists with the appropriate adjectives.

#### 3. Sentiment Analysis

##### System Prompt

You are an AI assistant, answer questions honestly, helpfully, and harmlessly.

##### User Prompt

Tell me about Abemaciclib.

#### 4. IrAE Detection

##### System Prompt

You are an expert Oncologist reviewing a note from the previous visit. Your task is to review the note step by step and answer the question using only a single number.

##### User Prompt

EHR Context:

Age: 68 years

Gender: Female

Cancer diagnosis: Stage IV non-small cell lung cancer (NSCLC)

PMH: hypertension

Prior cancer treatments: None

Current cancer treatments: Atezolizumab (started 3 weeks ago)

Current medication list: lisinopril, hydrochlorothiazide

Summary of most recent oncology visit (1 week ago): 68-year-old female with newly diagnosed stage IV NSCLC. She is on Atezolizumab monotherapy and tolerating treatment well. Will continue treatment as planned.

New complaint: Patient reports, "I've developed an itchy rash all over my body".

What is the likelihood this is an IrAE?

1 = Very Unlikely

2 = Unlikely

3 = Likely

4 = Very Likely

#### 5. IrAE Differential

##### System Prompt

You are an expert Oncologist reviewing a note from the previous visit. Your task is to review the note step by step and provide a Python list of diagnoses in order of probability.

##### User Prompt

EHR Context:

Age: 68 years

Gender: Female

Cancer diagnosis: Stage IV non-small cell lung cancer (NSCLC)

PMH: hypertension

Prior cancer treatments: None

Current cancer treatments: Atezolizumab (started 3 weeks ago)

Current medication list: lisinopril, hydrochlorothiazide

Summary of most recent oncology visit (1 week ago): 68-year-old female with newly diagnosed stage IV NSCLC. She is on Atezolizumab monotherapy and tolerating treatment well. Will continue treatment as planned.

New complaint: Patient reports, "I've developed an itchy rash all over my body".

Based on the information provided, what are the most likely diagnoses? Please provide your answer as a Python list, ordered from most likely to least likely.

#### 6. Drug Contraindications

##### System Prompt

You are a helpful AI assistant. Please provide the requested information accurately and concisely. You will be given multiple choice questions. You must only answer the questions with only the corresponding numbers for the answer.

##### User Prompt

Question: Which of the following drugs are contraindicated with Daunorubicin?

Choices:

1. Lomustine

2. Tetracycline

3. Mitoxantrone

4. Aldesleukin

Please provide the correct answer (1, 2, 3, or 4) only. Do not provide any other information.
