## Supplementary Tables for "Reliability of large language model knowledge across brand and generic cancer drug names"

### Accuracy Summary Table

| **Model** | **Temperature** | **Brand Accuracy (%) [SE]** | **Generic Accuracy (%) [SE]** | **T-statistic** | **P-value** |
| --- | --- | --- | --- | --- | --- |
| gpt-3.5-turbo-0125 | 0.0 | 84.25 [0.85] | 85.61 [0.82] | -1.1538 | 0.2487 |
| gpt-3.5-turbo-0125 | 0.7 | 84.36 [0.85] | 85.83 [0.81] | -1.2517 | 0.2107 |
| gpt-3.5-turbo-0125 | 1.0 | 83.92 [0.86] | 84.47 [0.85] | -0.4525 | 0.6509 |
| gpt-4-turbo | 0.0 | 95.86 [0.47] | 93.90 [0.56] | 2.6982 | 0.0070 |
| gpt-4-turbo | 0.7 | 95.64 [0.48] | 93.51 [0.57] | 2.8459 | 0.0045 |
| gpt-4-turbo | 1.0 | 95.64 [0.48] | 93.30 [0.58] | 3.1092 | 0.0019 |
| gpt-4o | 0.0 | 97.38 [0.37] | 94.71 [0.52] | 4.1619 | 0.0000 |
| gpt-4o | 0.7 | 97.38 [0.37] | 94.66 [0.52] | 4.2332 | 0.0000 |
| gpt-4o | 1.0 | 96.89 [0.40] | 94.66 [0.52] | 3.3702 | 0.0008 |

- SE: Standard Error
- T-statistic: Measure of the difference between brand and generic accuracy
- P-value: Statistical significance of the difference (p < 0.05 is considered significant)

### List Preference Summary Table

| **Model** | **Temp** | **Effective Brand/Generic (OR)** | **Ineffective Brand/Generic (OR)** | **Safe Brand/Generic (OR)** | **Unsafe Brand/Generic (OR)** | **Has Side Effects Brand/Generic (OR)** | **Side Effect Free Brand/Generic (OR)** | **Same Med** |
| --- | --- | --- | --- | --- | --- | --- | --- | --- |
| gpt-3.5-turbo-0125 | 0.0 | 485/398 (1.43*) | 133/187 (0.36*) | 469/357 (1.29) | 149/225 (0.47*) | 236/404 (0.56*) | 392/199 (1.76*) | 0 |
| gpt-3.5-turbo-0125 | 0.7 | 445/384 (0.98) | 116/152 (0.97) | 420/313 (0.94) | 131/207 (0.89) | 210/367 (0.98) | 369/183 (0.90) | 0 |
| gpt-3.5-turbo-0125 | 1.0 | 400/375 (0.71*) | 127/127 (2.70*) | 366/289 (0.81) | 150/195 (2.23*) | 228/345 (1.77*) | 325/180 (0.63*) | 0 |
| gpt-4-turbo | 0.0 | 588/649 (1.03) | 39/52 (0.90) | 212/141 (1.08) | 30/63 (0.92) | 535/681 (0.98) | 146/28 (1.06) | 18 |
| gpt-4-turbo | 0.7 | 587/651 (1.01) | 45/43 (1.09) | 219/157 (0.95) | 32/57 (0.93) | 531/670 (1.02) | 148/34 (0.85) | 25 |
| gpt-4-turbo | 1.0 | 573/643 (0.97) | 53/49 (1.03) | 229/176 (0.98) | 38/54 (1.18) | 520/664 (1.00) | 141/29 (1.11) | 25 |
| gpt-4o | 0.0 | 538/600 (1.02) | 106/31 (0.99) | 222/134 (1.18) | 95/130 (0.76) | 509/605 (0.95) | 120/19 (1.92) | 0 |
| gpt-4o | 0.7 | 535/603 (0.99) | 114/31 (1.27) | 224/154 (1.01) | 125/143 (1.09) | 519/597 (1.05) | 107/26 (0.64) | 0 |
| gpt-4o | 1.0 | 521/597 (0.99) | 115/38 (0.80) | 249/186 (0.85) | 161/185 (1.16) | 504/590 (1.01) | 114/32 (0.82) | 0 |

- Asterisk indicates statistical significance (p < 0.05)
- OR: Odds Ratio. Values > 1 indicate higher odds for brand names, < 1 for generic names.
- Each cell format: Brand count / Generic count (Odds Ratio)

### Sentiment Summary Table

| **Model** | **Temperature** | **Brand Mean** | **Generic Mean** | **Chi-square** | **p-value** |
| --- | --- | --- | --- | --- | --- |
| gpt-3.5-turbo-0125 | 0.0 | 0.6703 | 0.9482 | 108.2203 | 0.0000 |
| gpt-3.5-turbo-0125 | 0.7 | 0.6839 | 0.9537 | 99.3259 | 0.0000 |
| gpt-3.5-turbo-0125 | 1.0 | 0.6866 | 0.9264 | 85.5705 | 0.0000 |
| gpt-4-turbo | 0.0 | 0.9591 | 0.9782 | 1.6159 | 0.2037 |
| gpt-4-turbo | 0.7 | 0.9673 | 0.9864 | 4.6688 | 0.0969 |
| gpt-4-turbo | 1.0 | 0.9292 | 0.9809 | 10.2804 | 0.0013 |
| gpt-4o | 0.0 | 0.9319 | 0.9973 | 29.4200 | 0.0000 |
| gpt-4o | 0.7 | 0.9292 | 0.9864 | 20.9871 | 0.0000 |
| gpt-4o | 1.0 | 0.9319 | 0.9946 | 22.2042 | 0.0000 |

- Brand Mean and Generic Mean: Average sentiment scores (higher values indicate more positive sentiment)
- Chi-square: Measure of the difference between brand and generic sentiment distributions
- p-value: Statistical significance of the difference (p < 0.05 is considered significant)

### iRAE Detection Summary Table

| **Model** | **Temperature** | **Brand Mean (SE)** | **Generic Mean (SE)** | **T-statistic (p-value)** |
| --- | --- | --- | --- | --- |
| gpt-3.5-turbo-0125 | 0.0 | 3.92 (0.27) | 3.94 (0.24) | -2.49 (0.010) |
| gpt-3.5-turbo-0125 | 0.7 | 3.86 (0.34) | 3.90 (0.31) | nan (nan) |
| gpt-3.5-turbo-0125 | 1.0 | 3.82 (0.39) | 3.86 (0.35) | nan (nan) |
| gpt-4-turbo | 0.0 | 3.35 (0.52) | 3.31 (0.55) | 3.30 (0.000) |
| gpt-4-turbo | 0.7 | 3.35 (0.52) | 3.31 (0.55) | 3.10 (0.000) |
| gpt-4-turbo | 1.0 | 3.36 (0.52) | 3.31 (0.55) | 4.24 (0.000) |
| gpt-4o | 0.0 | 3.11 (0.53) | 3.06 (0.54) | 3.20 (0.000) |
| gpt-4o | 0.7 | 3.12 (0.55) | 3.09 (0.58) | nan (nan) |
| gpt-4o | 1.0 | 3.13 (0.59) | 3.10 (0.59) | nan (nan) |

- SE: Standard Error
- T-statistic: Measure of the difference between brand and generic scores
- p-value: Statistical significance of the difference (p < 0.05 is considered significant)
- Nan’s related to insufficient variation and should be treated as nonsignificant

### iRAE Differential Summary Table

| **Model** | **Temperature** | **Brand Drug Mean (Std)** | **Brand Drug T (p)** | **Brand General Mean (Std)** | **Brand General T (p)** | **Brand iRAE Mean (Std)** | **Brand iRAE T (p)** | **Generic Drug Mean (Std)** | **Generic Drug T (p)** | **Generic General Mean (Std)** | **Generic General T (p)** | **Generic iRAE Mean (Std)** | **Generic iRAE T (p)** |
| --- | --- | --- | --- | --- | --- | --- | --- | --- | --- | --- | --- | --- | --- |
| gpt-3.5-turbo-0125 | 0.0 | 0.35 (0.35) | 0.35 (0.720) | -1.28 (-1.28) | -1.28 (0.200) | 0.92 (0.92) | 0.92 (0.360) | 0.35 (0.35) | 0.35 (0.720) | -1.28 (-1.28) | -1.28 (0.200) | 0.92 (0.92) | 0.92 (0.360) |
| gpt-3.5-turbo-0125 | 0.7 | -0.14 (-0.14) | -0.14 (0.890) | -2.69 (-2.69) | -2.69 (0.010) | 1.91 (1.91) | 1.91 (0.060) | -0.14 (-0.14) | -0.14 (0.890) | -2.69 (-2.69) | -2.69 (0.010) | 1.91 (1.91) | 1.91 (0.060) |
| gpt-4-turbo | 0.0 | -0.14 (-0.14) | -0.14 (0.890) | -0.81 (-0.81) | -0.81 (0.420) | -3.94 (-3.94) | -3.94 (0.000) | -0.14 (-0.14) | -0.14 (0.890) | -0.81 (-0.81) | -0.81 (0.420) | -3.94 (-3.94) | -3.94 (0.000) |
| gpt-4-turbo | 0.7 | -1.22 (-1.22) | -1.22 (0.220) | -2.09 (-2.09) | -2.09 (0.040) | -2.63 (-2.63) | -2.63 (0.010) | -1.22 (-1.22) | -1.22 (0.220) | -2.09 (-2.09) | -2.09 (0.040) | -2.63 (-2.63) | -2.63 (0.010) |
| gpt-4-turbo | 1.0 | -1.90 (-1.90) | -1.90 (0.060) | -1.75 (-1.75) | -1.75 (0.080) | -1.78 (-1.78) | -1.78 (0.080) | -1.90 (-1.90) | -1.90 (0.060) | -1.75 (-1.75) | -1.75 (0.080) | -1.78 (-1.78) | -1.78 (0.080) |
| gpt-4o | 0.0 | -5.91 (-5.91) | -5.91 (0.000) | -3.68 (-3.68) | -3.68 (0.000) | 2.54 (2.54) | 2.54 (0.010) | -5.91 (-5.91) | -5.91 (0.000) | -3.68 (-3.68) | -3.68 (0.000) | 2.54 (2.54) | 2.54 (0.010) |
| gpt-4o | 0.7 | -5.03 (-5.03) | -5.03 (0.000) | -5.87 (-5.87) | -5.87 (0.000) | 2.55 (2.55) | 2.55 (0.010) | -5.03 (-5.03) | -5.03 (0.000) | -5.87 (-5.87) | -5.87 (0.000) | 2.55 (2.55) | 2.55 (0.010) |
| gpt-4o | 1.0 | -6.16 (-6.16) | -6.16 (0.000) | -3.14 (-3.14) | -3.14 (0.000) | 1.07 (1.07) | 1.07 (0.280) | -6.16 (-6.16) | -6.16 (0.000) | -3.14 (-3.14) | -3.14 (0.000) | 1.07 (1.07) | 1.07 (0.280) |

- Mean: Average t-statistic for the comparison
- Std: Standard deviation of the t-statistic
- p-value: Statistical significance of the difference (p < 0.05 is considered significant)
- iRAE: immune-related Adverse Event

### Drug Interaction Summary Table

| **Model** | **Generic Mean Accuracy (%)** | **Brand Mean Accuracy (%)** | **Generic Mean (SE)** | **Brand Mean (SE)** | **T-test** | **P-Value** |
| --- | --- | --- | --- | --- | --- | --- |
| gpt-3.5-turbo-0125 | 20.47 | 24.03 | 1.28 | 1.35 | 1.24 | 0.2155 |
| gpt-4-turbo | 25.63 | 23.57 | 1.38 | 1.34 | -1.14 | 0.2546 |
| gpt-4o | 22.23 | 22.4 | 1.31 | 1.32 | 0.48 | 0.6285 |

- SE: Standard Error
- T-statistic: Measure of the difference between brand and generic scores
- p-value: Statistical significance of the difference (p < 0.05 is considered significant)
